# Clinical Evidence Supporting FDA Clearance of First-of-a-Kind Therapeutic Devices via the De Novo Pathway Between 2011 and 2019

**DOI:** 10.1101/2020.04.23.20077164

**Authors:** James L. Johnston, Sanket S. Dhruva, Joseph S. Ross, Vinay K. Rathi

**Affiliations:** Yale School of Medicine, New Haven, CT; San Francisco Veterans Affairs Medical Center, San Francisco, CA; Department of Medicine, University of California, San Francisco School of Medicine, San Francisco, CA; Center for Outcomes Research and Evaluation (CORE), Yale-New Haven Hospital, New Haven, CT; Department of Health Policy and Management, Yale School of Public Health, New Haven, CT; Section of General Internal Medicine, Department of Internal Medicine, Yale School of Medicine, New Haven, CT; Department of Otolaryngology - Head and Neck Surgery, Massachusetts Eye and Ear Infirmary, Boston, MA; Harvard Business School, Boston, MA

**Author notes:** **Correspondence to:** Vinay K. Rathi, MD, Department of Otolaryngology - Head and Neck Surgery, Massachusetts Eye and Ear Infirmary, 243 Charles Street, Boston, MA 02114.

## Abstract

**Importance:** In recent years, the US Food and Drug Administration (FDA) and manufacturers have increasingly sought to expedite patient access to first-of-a-kind devices via the De Novo premarket review pathway. Understanding the strength of clinical evidence supporting FDA clearance through this pathway can help guide clinical adoption of novel devices and ongoing regulatory development of the postmarket surveillance infrastructure.

**Objective:** Our primary objective was to characterize the strength of clinical evidence supporting FDA clearance of therapeutic De Novo devices. Key secondary objectives were 1) characterization of FDA post-marketing requirements for De Novo devices and 2) study of the use of these devices as the basis for devices subsequently cleared via the 510(k) process.

**Design:** Retrospective cross-sectional analysis

**Setting:** Publicly available online FDA databases, including the De Novo database, the 510(k) clearance database, the 522 Post Market Surveillance database, and the Recalls of Medical Devices database

**Participants:** All moderate-risk therapeutic devices cleared via the De Novo pathway between January 1, 2011, and December 31, 2019.

**Main Outcome Measures:** (1) proportion of De Novo devices cleared based on evidence from a pivotal clinical study, (2) proportion of pivotal study primary effectiveness endpoints that were met, (3) proportion of De Novo devices subject to FDA-required postmarket studies, and (4) proportion of De Novo devices serving as the basis for at least one subsequently cleared 510(k) device (i.e., new models or competitor products).

**Results:** There were 63 (of 65; 96.9%) moderate-risk therapeutic devices cleared by FDA via the De Novo pathway between 2011 and 2019 for which decision summary documentation was publicly available. Of the 63 devices, 51 (81.0%) were supported by pivotal clinical studies (n=54 studies); the remainder (n= 12; 19.0%) were not supported by a pivotal clinical study. The majority of pivotal studies were randomized (57.4%), multi-armed (61.1%), and used an active (25.9%) or sham (35.2%) comparator arm; 17 (31.5%) failed to meet at least one primary effectiveness endpoint. Among the 63 devices cleared via the De Novo pathway, one (1.6%) was subject to an FDA-required posttmarket study and 32 (47.8%) served as a predicate device for new models or competitor devices subsequently cleared through the 510(k) process.

**Conclusions:** Between 2011 and 2019, the FDA cleared the majority of first-of-a-kind moderate-risk therapeutic devices via the De Novo pathway based on premarket evidence from pivotal clinical studies. However, 43% of devices were cleared without clinical evidence from pivotal studies or based on pivotal studies that failed to meet at least one primary effectiveness endpoint. The FDA rarely required postmarket studies of these devices, which often served as the basis for new models and competitor products subsequently cleared via the 510(k) process.

**KEY POINTS:** *Question:* What is the strength of premarket clinical evidence supporting FDA clearance of first-of-a-kind therapeutic devices via the De Novo review pathway?

*Findings:* In this retrospective cross-sectional analysis of 63 devices, 43% were cleared without supporting premarket pivotal studies or based on pivotal studies that failed to meet at least one primary effectiveness endpoint. The FDA rarely required postmarket studies of therapeutic De Novo devices, which often served as the basis for new models and competitor products subsequently cleared via the 510(k) process.

*Meaning:* The FDA often clears first-of-a-kind therapeutic devices via the De Novo pathway despite limited clinical evidence of effectiveness.

## INTRODUCTION

Under the Medical Device Regulation Act of 1976,^1^ the US Food and Drug Administration (FDA) regulates medical devices using a three-tiered risk classification system: class I (low-risk devices, such as bandages), class II (moderate-risk devices, such as glucometers), and class III devices (high-risk devices, such as prosthetic heart valves). Approximately 90% of devices subject to FDA premarket review are classified as moderate-risk.^2^ Moderate-risk devices are primarily regulated through the 510(k) pathway, which requires manufacturers to demonstrate their device is “substantially equivalent” in intended use and technological specifications (with allowable exceptions) to at least one previously FDA-cleared device, known as a “predicate device.”^3^ Fewer than 10% of 510(k) devices are cleared based on supporting premarket clinical evidence; the safety and effectiveness of these devices is presumed based on the evaluation of the predicate device.

The 510(k) pathway was not designed to regulate first-of-a-kind devices; as a result, the FDA was historically required to automatically designate all novel technologies as high-risk, even for devices that presumably conferred lower levels of risk to patients. These technologies were subject to regulation via the Premarket Approval pathway for high-risk devices, which requires clinical evidence for approval and is therefore the longest and most costly route to market. In an effort to reduce barriers to technological innovation and patient access, US Congress established the De Novo pathway under the Food and Drug Administration Modernization Act of 1997.^4^ The De Novo pathway permits manufacturers to establish and market first-of-a-kind low- or moderate risk devices, which may serve as predicates supporting clearance of subsequent 510(k) devices.

Device manufacturers can apply for device clearance via the De Novo pathway following FDA denial of preceding 510(k) applications due to lack of substantial equivalence or (as of 2012) directly when devices have no potential predicates. De Novo pathway applications require several key components, including the recommended risk-classification for the device, the probable benefits and risks accompanying device use, and non-clinical data such as bench performance testing. If non-clinical studies are deemed insufficient to provide reasonable assurance of device safety and effectiveness, the FDA requires clinical data for clearance.^5^

The De Novo pathway has served as a path to market for a variety of devices, such as the single-lead electrocardiogram feature of the Apple© Watch and treatments (e.g., embolization and high intensity therapeutic ultrasound devices) for benign prostatic hyperplasia.^5^ In recent years, manufacturers have increasingly sought to market first-of-a-kind devices via the De Novo pathway. Whereas the FDA cleared 65 devices via the De Novo pathway between 1997 and 2012, the agency cleared 165 devices through this pathway between 2013 and 2018.^6^ The FDA anticipates that the importance of the De Novo pathway will continue to grow as the agency seeks to expedite patient access to novel devices relative to international markets.^6^

Given the increasing use of this pathway for novel medical devices, patients, physicians, and policymakers should understand the evidence supporting De Novo clearance. This evidence has important implications for clinical practice, as premarket studies inform both the clinical adoption of De Novo devices as well as subsequent FDA regulation of new models and competitor products, including the need for postmarket surveillance. We therefore sought to systematically characterize the premarket pivotal clinical studies supporting FDA clearance of therapeutic devices cleared via the De Novo Pathway and FDA postmarket experience with these devices.

## METHODS

### Device Cohort

We conducted a retrospective cross-sectional analysis of therapeutic devices cleared through the De Novo pathway between January 1, 2011, and December 31, 2019, using the FDA De Novo database (**Figure 1**).^7^ We restricted our analysis to devices cleared after 2010 because the FDA began releasing documents summarizing the premarket evidence supporting clearance during this year; devices without publicly available summary documents were excluded from analysis.^8^ We chose to focus on therapeutic devices because these technologies are typically evaluated based on clinical safety and effectiveness data rather than concordance with reference standards (as is often the case for diagnostic devices).

**Figure 1.**
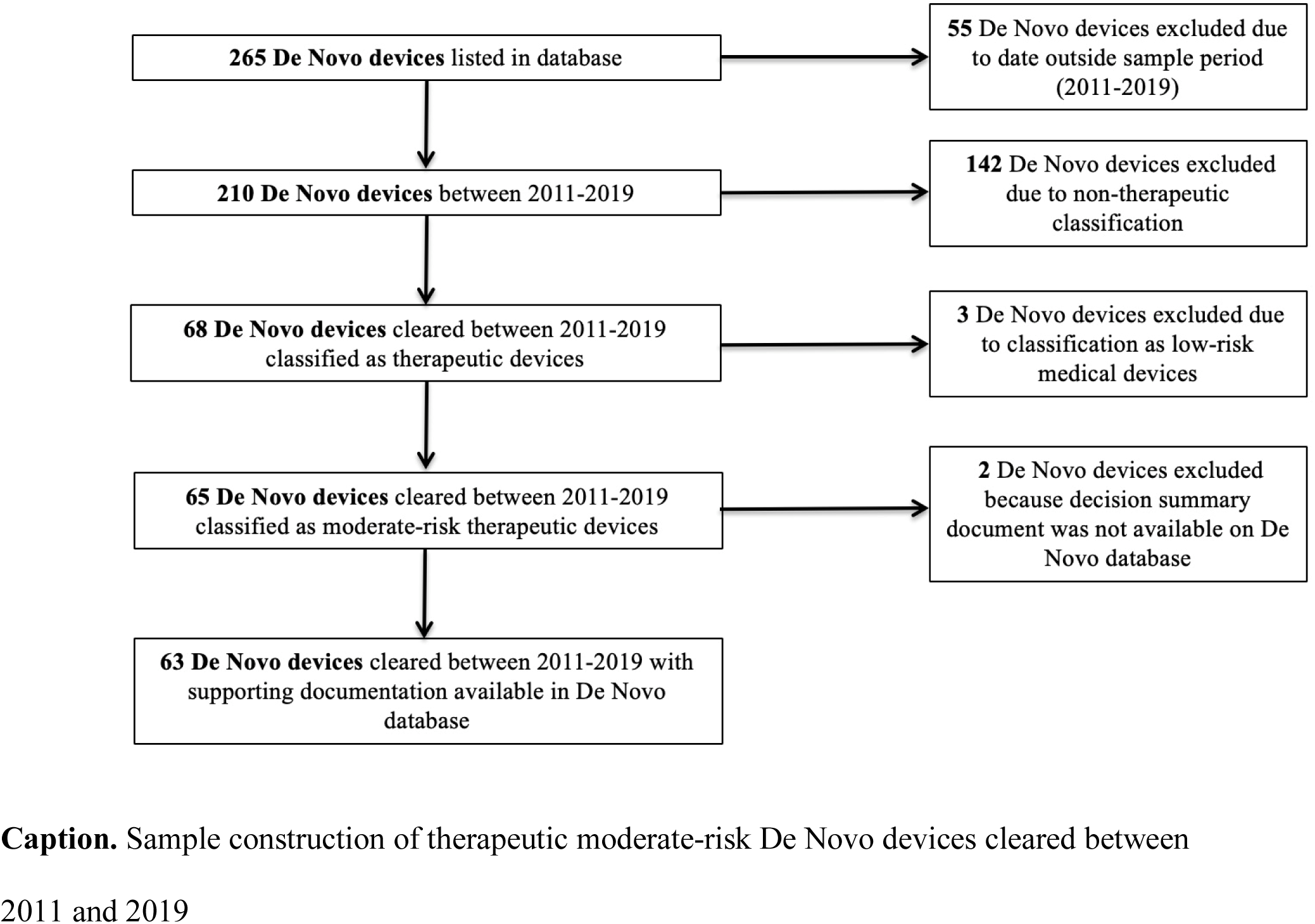
Sample Construction of Therapeutic Moderate-Risk De Novo Devices Cleared Between 2011 and 2019

Using FDA-designed product codes,^9^ we classified each De Novo device as either therapeutic or non-therapeutic (e.g., diagnostic tests or procedural equipment). For each therapeutic device, we determined the clearance year, FDA-designated medical specialty, and whether the manufacturer submitted the marketing application directly via the De Novo pathway or following a denied 510(k) application. We additionally extracted whether the FDA classified the device as implantable (yes/no) and life-sustaining (yes/no).

### Premarket Study Evidence: Pivotal Studies

There are two types of premarket clinical studies for medical devices – non-pivotal studies (e.g., small-scale feasibility studies) and pivotal studies, which generally serve as the primary basis for device clearance.^10^ Using previously described methods, we reviewed publicly available documents within the FDA De Novo database to identify all pivotal clinical studies supporting clearance of devices in our sample.^11^ We categorized each pivotal study by the following characteristics: total number of patients enrolled, total number of patients enrolled in treatment device group, comparator type (active/sham/pre-post comparison/historical/none), randomization (yes/no), blinding (yes/no), study arms (single/multiple), study location (US/Outside US/mixed), number of study centers (single-center/multi-center), and duration of longest primary effectiveness endpoint follow-up. For each study, we categorized all primary endpoints by the following characteristics: outcome type (safety/effectiveness), effectiveness endpoint type (clinical outcome/clinical scale/surrogate marker), and endpoint met (yes/no/not specified).^11^

### Postmarket Experience: Postmarketing Requirements, Recalls, and Subsequent 510(k) Devices

We characterized FDA postmarket experience with devices in our sample by several means. First, we identified all FDA-required postmarket studies of these devices. FDA-required postmarket studies are intended to complement premarket understanding of device benefits and risks. To identify postmarket study requirements, we queried the 522 Studies database on March 22, 2020 by device application number.^12^ The FDA may order 522 Studies for moderate- or high-risk devices when device failure would be reasonably likely to cause significant health problems. We categorized each required postmarket study by purpose (evaluation of safety/evaluation of effectiveness).

Second, we extracted the recall history of the De Novo devices in our sample using the FDA Recalls of Medical Devices database. We queried this database on March 1, 2020, using device application numbers to determine if a device was recalled and, if applicable, the FDA-designated recall class (Class I – highest risk/Class II – moderate risk/Class III – low risk).^13^

Third, we identified all 510(k) clearances of new models and competitor products based on De Novo devices in our sample. For each device, we queried the FDA 510(k) database on March 13, 2020,^14^ using FDA-designated product codes. We collected the dates of first clearance for: (1) new models marketed by De Novo manufacturers and; (2) first competitor devices marketed by other manufacturers. We also collected the number of 510(k) clearances for each device and market lifespan (in days) as of March 13, 2020.

### Statistical Analysis

We used descriptive statistics to characterize therapeutic De Novo devices, premarket pivotal studies and primary endpoints, postmarket study requirements, and 510(k) device lineages. For each De Novo device linked to a subsequent 510(k) device, we calculated median time to first 510(k) device, median time to first competitor device clearance, and median number of subsequent 510(k) clearances per year. All analyses were performed in Microsoft Excel (Microsoft Corporation; Redmond, Washington, USA).

## RESULTS

### Device Cohort

Between January 1, 2011, and December 31, 2019, the FDA cleared 210 first-of-a-kind devices through the De Novo pathway (**Figure 1**). Among these 210 devices, 68 (32.4%) were therapeutic. Of those 68 devices, 65 (95.6%) were FDA-classified as moderate-risk. Of these 65 devices, decision summary documents were available in the FDA’s De Novo database for 63 (96.9%).

Among these 63 first-of-a-kind moderate-risk therapeutic devices, the FDA classified zero (0.0%) as life-sustaining and 10 (15.9%) as implantable (**Table 1**). The most commonly represented medical specialties were neurology (18 of 63: 28.6%), gastroenterology and urology (13 of 63: 20.6%), and general and plastic surgery (10 of 63: 15.9%). Nearly three-quarters (48 of 63: 76.2%) were cleared through the direct De Novo pathway, with the remainder (15 of 63: 23.8%) cleared following FDA denial of a preceding 510(k) application.

**Table 1.** First-of-a-kind moderate-risk therapeutic devices cleared by the US Food and Drug Administration through the De Novo Pathway Between 2011 and 2019

|  | No. (%)<br>(n = 63) |
| --- | --- |
| Approval Year |  |
| 2011-2014 | 20 (31.7) |
| 2015-2019 | 43 (68.3) |
| Application Type |  |
| Direct De Novo | 48 (76.2) |
| Post 510(k) Denial | 15 (23.8) |
| Specialty |  |
| Neurology | 18 (28.6) |
| Gastroenterology and Urology | 13 (20.6) |
| General & Plastic Surgery | 10 (15.9) |
| Otolaryngology | 5 (7.9) |
| Ophthalmology | 4 (6.3) |
| Anesthesiology | 4 (6.3) |
| Cardiovascular | 3 (4.8) |
| Other | 6 (9.5) |
| Review Type<sup>a</sup> |  |
| Standard | 62 (98.4) |
| Expedited | 1 (1.6) |
| Implantable |  |
| Yes | 10 (15.9) |
| No | 53 (84.1) |
| Life Sustaining |  |
| Yes | 0 (0) |
| No | 63 (100) |
| FDA Required Postmarket Study |  |
| Yes | 1 (1.6) |
| No | 62 (98.4) |
| Highest Recall Class |  |
| Class I | 0 |
| Class II | 6 (9.5) |
| Class III | 0 |
| No Recall | 57 (90.5) |

<sup>a</sup>Expedited indicates review via the FDA Priority Review Program, which was intended to facilitate patient access to therapies addressing unmet medical needs in the treatment of serious or life-threatening conditions

### Premarket Study Evidence: Pivotal Studies

Among the 63 moderate-risk, therapeutic medical devices, 12 (19.0%) devices were not supported by a pivotal study; instead, these devices were cleared on the basis of non-pivotal (feasibility) studies, literature review of similar devices, or postmarket data from other localities such as the European Union (**Table 2**). Examples of De Novo devices that were not supported by premarket pivotal studies include a transesophageal core temperature cooling device, a cerebrospinal fluid drainage system for patients with post-operative neurologic deficits, and a fertility planning software.

**Table 2.** Clinical Evidence Supporting De Novo devices not supported by a premarket pivotal study

| Application Number | Device Manufacturer | Device Name | Clearance Date | Device Specialty | FDA<sup>a</sup> Cleared Clinical Indication | Clinical Evidence Submitted |
| --- | --- | --- | --- | --- | --- | --- |
| EN180058 | Tandem Diabetes Care, Inc. | t:slim X2 insulin pump with interoperable technology | 2/14/2019 | Clinical Chemistry | Subcutaneous delivery of insulin, at set and variable rates, for the management of diabetes mellitus in persons requiring insulin. | No clinical data included in decision summary |
| EN180013 | KCI USA, Inc. | Prevena 125 and Prevena Plus 125 Therapy Units | 4/19/2019 | General & Plastic Surgery | Aid in reducing the incidence of post-operative seroma and – in high-risk patients – the risk of superficial surgical site infections | Systematic review and meta-analysis of 16 clinical studies evaluating the device, including 2 previously unpublished manufacturer-sponsored trials. |
| EN170052 | Natural Cycles Nordic AB | Natural Cycles | 8/10/2018 | Obstetrics/ Gynecology | Fertility monitoring software for women > 18 years to help plan or prevent pregnancy | Real-world postmarket data from 37 countries outside the United States |
| EN170044 | Thornhill Research, Inc. | ClearMate | 3/14/2019 | Anesthesiology | Adjunctive treatment to be used by emergency department clinicians for patients suffering from carbon monoxide poisoning. | - Literature review of 5 published studies evaluating the device.<br>- European study evaluating device for different indication used to support device safety |
| EN170015 | Wilson-Cook Medical, Inc | Hemospray Endoscopic Hemostat | 5/7/2018 | General & Plastic Surgery | Hemostasis of non-variceal gastrointestinal bleeding. | - One pilot premarket study<br>- Two postmarket studies outside the United States |
| EN160040 | Biosphere Medical S.A. | Embosphere Microspheres | 6/21/2017 | Gastroenterology & Urology | Embolization of arteriovenous malformations, hypervascular tumors, including symptomatic uterine fibroids, and prostatic arteries for symptomatic benign prostatic hyperplasia . | - Review of literature supporting device use for intended indication.<br>- Partial results from 3 ongoing phase II trials<br>- Database from outside the United States (400+ patients) |
| EN140018 | Advanced Cooling Therapy, LLC | Esophageal Cooling Device | 6/23/2015 | Cardiovascular | 1. Control patient temperature<br>control patient temperature, and<br>2. Provide gastric decompression and suctioning | Outside the United States postmarket clinical data from 16 patients |
| EN130016 | Revmedx, Inc. | XSTAT | 4/3/2014 | General & Plastic Surgery | Temporary (maximum 4 hours) control of bleeding from junctional wounds in the groin or axilla not amenable to tourniquet application in adults and adolescents. Intended for battlefield use. | No clinical data |
| EN120017 | Medtronic Neurosurgery | Medtronic DUET External Drainage and Monitoring System | 8/22/2014 | Neurology | Temporary drainage and monitoring of cerebrospinal fluid (CSF) flow from the lumbar subarachnoid space in:<br>1. Patients undergoing open descending thoracic aortic aneurysm (open TAA) or open descending thoraco-abdominal aortic aneurysm (open TAAA) repair surgery.<br>2. Patients post TAA/TAAA repair that become symptomatic with neurological deficit such as paraplegia. | Review of 5 published studies evaluating CSF draining apparatuses with similar characteristics to the device (n=5) for device indications |
| EN100025 | Office Research in Clinical Amplification | Widexlink in Clear Series Hearing Aids | 3/31/2011 | Ear Nose Throat | Auditory amplification for individuals with a full range of hearing loss severity (from slight (16 to 25 dB HL) to profound (90+ dB HL)) and all hearing loss configurations. | FDA-requested pilot study (n=12 patients) |
| EN080015 | Numed, Inc. | Nucleus-X PTV Catheter | 6/11/2012 | Cardiovascular | Balloon aortic valvuloplasty for the treatment of aortic valve stenosis. | Review of literature (n=3 studies) on worldwide experience with balloon aortic valvuloplasty, including studies evaluating the Nucleus family of catheters |
| EN170086 | Allergan | TrueTear Intranasal Tear Neurostimulator | 5/17/2018 | Ophthalmology | Provides temporary increase in tear production during neurostimulation to improve dry eye symptoms in adult patients with severe dry eye symptoms. | De Novo clearance was for expanded indications from another De Novo device in the analyzed cohort. Clearance was supported by additional analysis of pivotal trials supporting first De Novo device, evaluating device effectiveness at symptom relief, but no new pivotal trials supported new De Novo clearance |

<sup>a</sup>FDA denotes the US Food and Drug Administration

In total, we identified 54 pivotal studies supporting the clearance of 51 (of 63; 81.0%) devices (**Table 3**). Among pivotal studies, 31 (57.4%) were randomized, 23 (42.6%) were blinded, 41 (75.9%) were multi-center, and 33 (61.1%) were multi-armed. Among multi-armed studies, 14 (42.4%) used an active comparator and 19 (57.6%) used a sham comparator. Among single-armed studies, 8 (38.1%) used participant pre/post comparisons, 11 (52.4%) used no comparator, and 2 (9.5%) used a historical comparator (**Table 3)**. Median study enrollment for all pivotal studies was 112.5 patients (IQR: 73.5-187). For multi-armed pivotal studies, median study enrollment was 150 (IQR: 100-206) patients and the median treatment group enrollment was 89 (IQR: 49-118.5) patients.

**Table 3.**
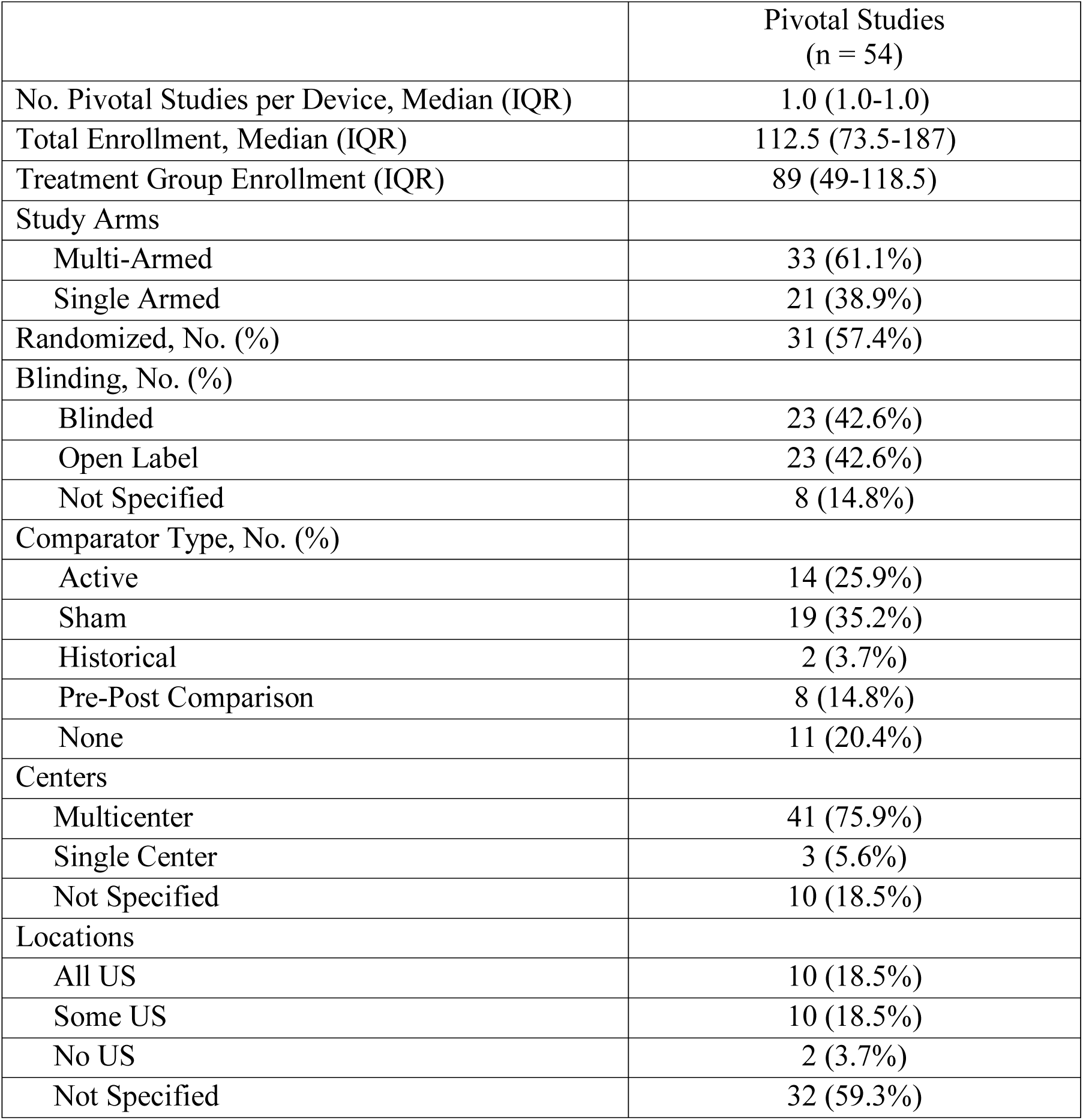
Characteristics of Pivotal Studies Supporting US Food and Drug Administration Clearance of De Novo Classification Therapeutic Medical Devices Between 2011 and 2019

|  | Pivotal Studies<br>(n = 54) |
| --- | --- |
| No. Pivotal Studies per Device, Median (IQR) | 1.0 (1.0-1.0) |
| Total Enrollment, Median (IQR) | 112.5 (73.5-187) |
| Treatment Group Enrollment (IQR) | 89 (49-118.5) |
| Study Arms |  |
| Multi-Armed | 33 (61.1%) |
| Single Armed | 21 (38.9%) |
| Randomized, No. (%) | 31 (57.4%) |
| Blinding, No. (%) |  |
| Blinded | 23 (42.6%) |
| Open Label | 23 (42.6%) |
| Not Specified | 8 (14.8%) |
| Comparator Type, No. (%) |  |
| Active | 14 (25.9%) |
| Sham | 19 (35.2%) |
| Historical | 2 (3.7%) |
| Pre-Post Comparison | 8 (14.8%) |
| None | 11 (20.4%) |
| Centers |  |
| Multicenter | 41 (75.9%) |
| Single Center | 3 (5.6%) |
| Not Specified | 10 (18.5%) |
| Locations |  |
| All US | 10 (18.5%) |
| Some US | 10 (18.5%) |
| No US | 2 (3.7%) |
| Not Specified | 32 (59.3%) |

We identified 60 primary effectiveness endpoints within these 54 pivotal studies; FDA documents specified no primary endpoint for five studies supporting five devices (**Table 4**). Of those 60 primary effectiveness endpoints, 36 (60.0%) evaluated clinical outcomes (e.g., reduction in hemhorrhoids rate during child delivery), 17 (28.3%) were clinical scales (e.g., reduction in Reflux Symptom Index), and 7 (11.7%) were surrogate markers of disease (e.g., increase in number of active Meibomian glands). The median duration of longest primary effectiveness endpoint follow up was 1.0 months (IQR: 0.0-3.0 months) for non-implantable devices and 9.5 months (IQR: 3.5-12.0 months) for implantable devices. All 27 (100.0%) primary safety endpoints were met.

**Table 4.** Characteristics of Primary Effectivess Endpoints of Pivotal Studies Supporting US Food and Drug Administration Clearance of De Novo Classification Therapeutic Medical Devices Between 2011 and 2019

|  |  |
| --- | --- |
|  | **Pivotal Premarket**<br>**Effectiveness**<br>**Endpoints**<br>**(n = 60)<sup>a</sup>** |
| **Endpoint Type, No. (%)** |  |
| Clinical Endpoint | 36 (60.0%) |
| Clinical Scale | 17 (28.3%) |
| Surrogate Marker | 7 (11.7%) |
| **Median Duration of Longest**<br>**Follow-Up (months) (IQR)** |  |
| Overall | 1.0 (0.0 - 6.0) |
| Non-Implantable | 1.0 (0.0 - 3.0) |
| Implantable | 9.5 (3.5 - 12.0) |
| **Primary Endpoint(s) Met** |  |
| Yes | 40 (66.7%) |
| No | 18 (30.0%) |
| Endpoint(s) Success not Evaluated | 2 (3.3%) |

**Notes:** FDA=Food and Drug Administration; IQR=Interquartile Range
<sup>a</sup> Does not include 5 pivotal studies supporting 5 devices without a primary effectiveness endpoint

Among primary effectiveness endpoints, 66.7% (n=40) were met, 30.0% (n=18) were not met, and 3.3% (n=2) did not have their success/failure disclosed within FDA documents. In total, 17 (of 54; 31.5%) pivotal studies supporting 15 (of 63; 23.8%) devices failed at least one primary effectiveness endpoint. FDA reasons for clearing these devices included the success of a co-primary endpoint, post hoc analyses suggesting benefit, and lower risk profile than therapeutic alternatives (e.g., pharmaceutical agents) (**Table 5**). Examples of De Novo devices that were supported by pivotal studies that failed primary effectiveness endpoints include a transcutaneous vagal nerve stimulator for migraine prophylaxis, treatment for urinary incontence, and embolic protection system used during transcather aortic valve replacement procedures.

**Table 5.**
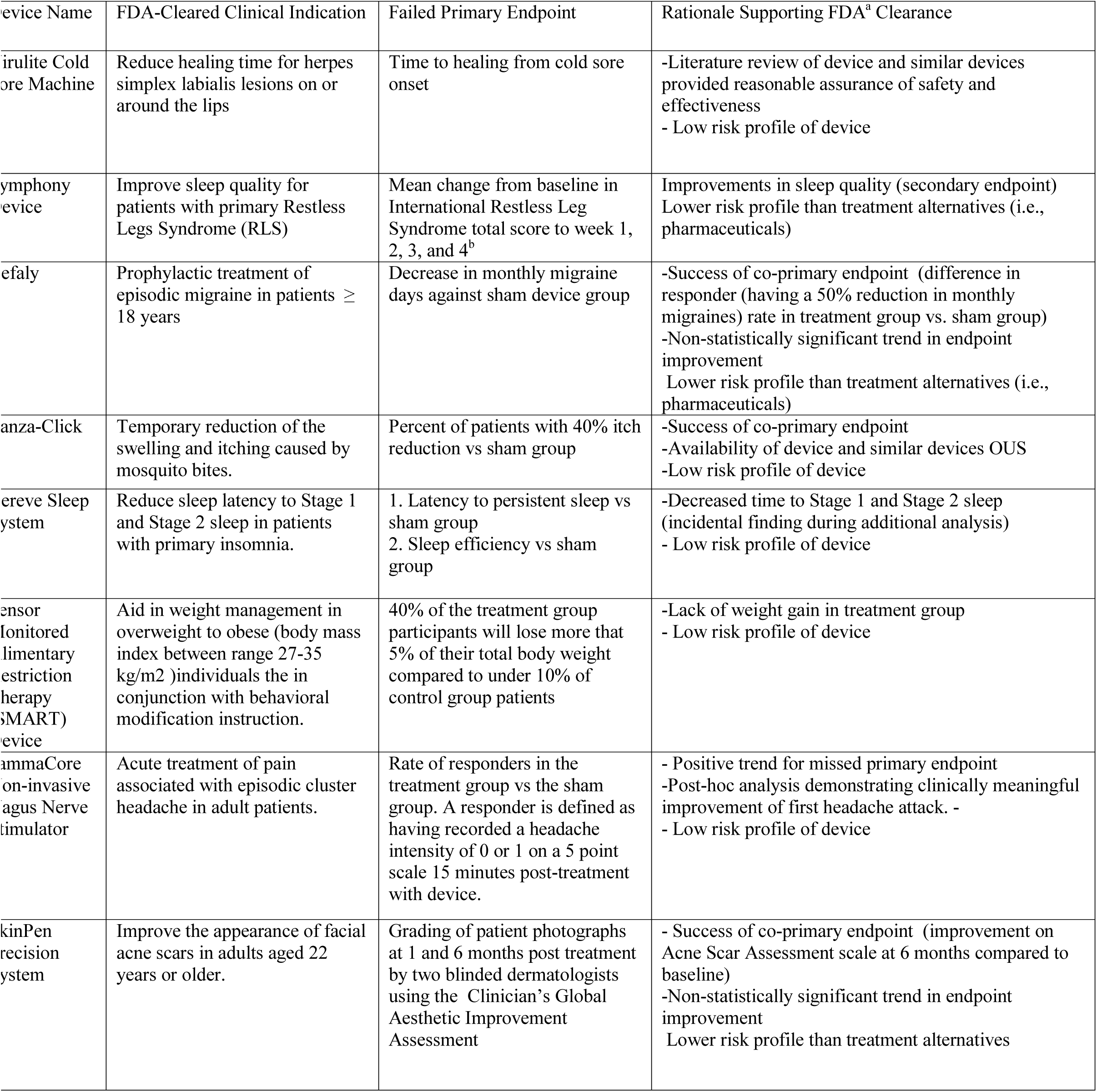
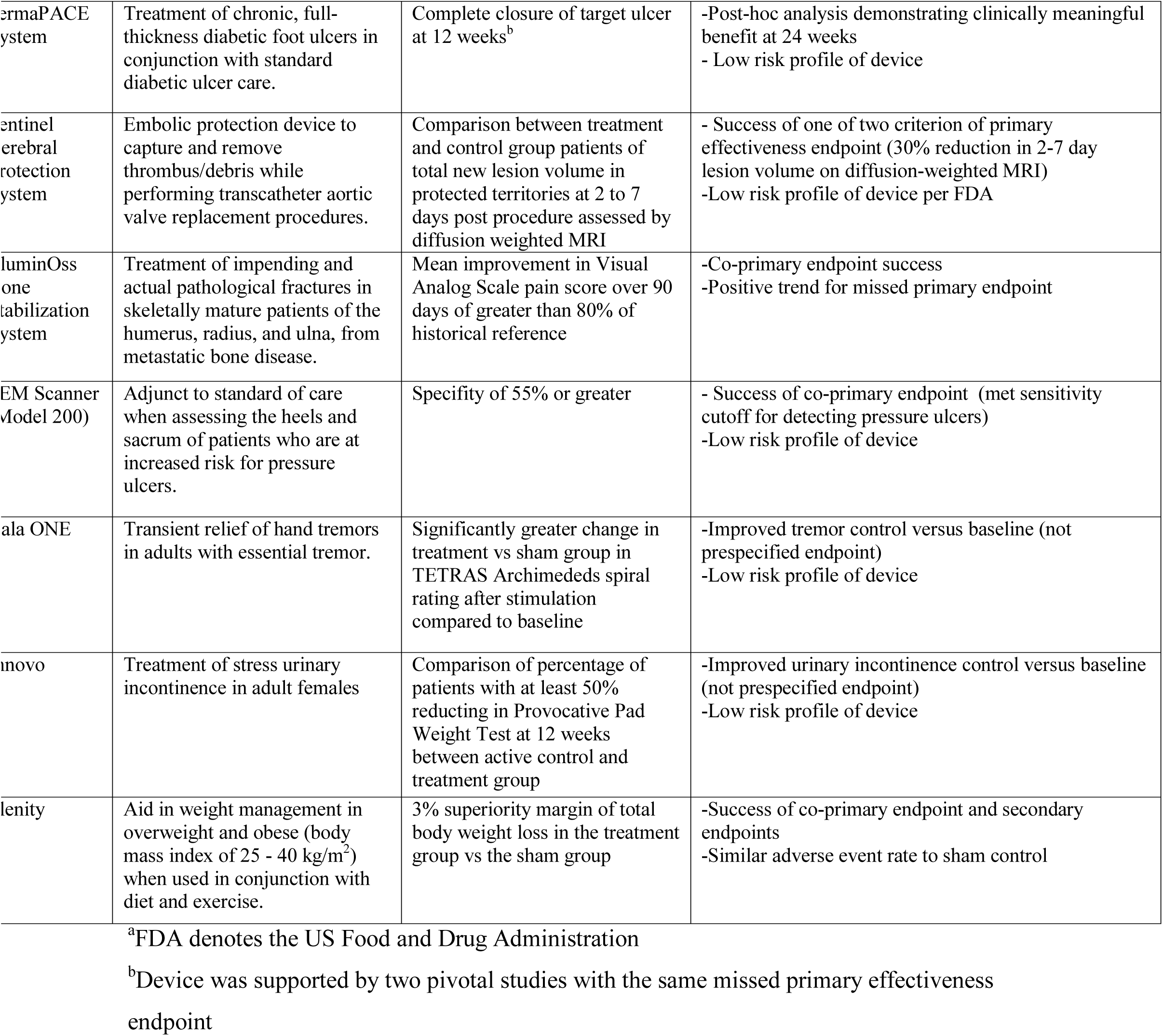
FDA Rationale for Clearance of De Novo Devices with one or more missed primary effectiveness endpoints in supporting pivotal studies

### Postmarket Experience: Postmarketing Requirements and Recalls

Between 2011 and 2019, the FDA required a postmarket study for one (of 63; 1.6%) therapeutic De Novo device. This device was a powered exoskeleton indicated to help patients with weakened or paralyzed limbs ambulate. This study was intended to examine the long-term safety of the device because failure could result in fall-related injuries. This device was subsequently recalled due to postmarket reports of patients suffering injury to their tibias and fibulas.

The vast majority (n=57 of 63; 90.5%) of De Novo devices were not recalled. Six devices (of 63: 9.5%) underwent class II (i.e., moderate-risk) recalls. Three devices were recalled due to shipping problems or improper device labeling. One device - an endoscopic hemostat for gastrointestinal bleeding - was recalled due to a malfunction with the potential to delay hemostasis. Two devices - a robotic system for benign prostatic hyperplasia and a localized heating device for chronic eyelid cysts - were recalled due to component issues that resulted in device failures.

### Postmarket Experience: Subsequent 510(k) Devices

Thirty-two (of 63: 50.8%) moderate-risk therapeutic De Novo devices served as the predicate for at least one device subsequently cleared via the 510(k) process (i.e., 510[k] devices). Among these De Novo devices, the median number of 510(k) devices was 2.5 (IQR: 1-5) and the median number of 510(k) devices per year was 0.74 (IQR: 0.54-1.14). The median time from initial De Novo clearance to first 510(k) device clearance was 202 (IQR: 159-302) days.

Sixteen (of 63; 25.4%) De Novo devices served as the basis for at least one device from a competing manufacturer. For these 16 devices, the median time from initial De Novo clearance to first competing manufacturer 510(k) device was 364 (IQR: 184.5-632.0) days. The competitor product served as the first 510(k) device for the majority (n=11 of 16; 68.8%) of these devices.

Eleven devices served as the basis for 510(k) devices marketed by both the De Novo device manufacturer and and a competing device manufacturer. For these devices, the median time to first competing manufacturer 510(k) device was 426 (IQR: 161.5-678.5) days after De Novo clearance and the median time to first De Novo manufacturer 510(k) device was 439 (IQR: 220-552) days.

## DISCUSSION

We found substantial variation in the strength of evidence supporting first-of-a-kind therapeutic medical devices cleared via the De Novo pathway between 2011 and 2019. The majority of devices were cleared based on evidence from pivotal clinical studies. Among pivotal studies, most were randomized, multi-armed studies with an active or sham comparator. However, 43% of devices were not evaluated through pivotal studies or supported by pivotal studies with at least one failed primary effectiveness endpoint

Despite these potential limitations in premarket evidence, the FDA required only one device – a powered exoskeleton – to undergo postmarket study. This device was subsequently recalled after causing lower extremity injuries. Nonetheless, overall few devices were recalled. These safety findings are reassuring, as nearly half of De Novo devices in our cohort served as a predicate device for a subsequent 510(k) device, including nearly a quarter that served as a predicate for a competitor product.

To our knowledge, our study is the first to systematically review medical devices cleared via the FDA De Novo pathway. In contrast to the oft scrutinized 510(k) process,^2,15^ the De Novo pathway typically requires manufacturers to submit clinical evidence of device safety and effectiveness. When pivotal studies are not required, the FDA often considers alternative clinical data sources, although these may be limited by biases due to selective publication or the challenges of detecting postmarket safety signals.^16,17^ When pivotal studies are required, premarket clinical evidence is generally less rigorous than that required for approval of high-risk devices via the Premarket Approval pathway.^11,18^ Less stringent evidentiary requirements are consistent with the original FDA aim of reducing barriers to market for these comparatively lower-risk devices, but have significant implications for patients and clinicians.

Much variability in the strength of premarket evidence supporting De Novo devices may be due to the diverse range of regulated technologies and indications for use. For instance, the evidentiary bar for a software based weight management aid might be reasonably lower than that for a balloon valvuloplasty catheter for treating aortic stenosis; both devices were cleared via the De Novo pathway. However, our findings reveal that the FDA often exercises regulatory disecretion when devices fail to acheive primary effectiveness endpoints. For such devices, the FDA determines probable benefit based on post-hoc analyses, alternative clinical data sources, and comparison to therapeutic alternatives. As a result, the analyses supporting these determinations may suffer from selection bias, inadequate statistical power, or other potential limiations.^19,20^

Though we found few recalls of De Novo devices, patients, physicians, and payors would benefit from more definitive evidence of device effectiveness. For example, in June 2017, the FDA cleared a cerebral embolic protection device indicated for use during transcatheter aortic valve replacement (TAVR) via the De Novo pathway, despite the fact that the supporting premarket pivotal study failed to meet its primary endpoint (reduction in new lesion volume within protected brain territories) by one of two preset criterion. The Centers for Medicare and Medicaid (CMS) services subsequently authorized supplemental payment to facilitate device utilization.^21,22^ Recent study of postmarket experience reveals that the device was rapidly adopted into clinical practice, but did not result in significant reduction in post-TAVR stroke or transient ischemic attack.^21,23^ This saga illustrates the need for more rigorous evidentiary standards, which could help minimize clinical opportunity costs and wasteful healthcare spending that result from the adoption of ineffective devices.

De Novo device manufacturers may be reluctant to invest in more rigorous clinical studies under the current regulatory framework for moderate-risk devices. At present, these manufacturers are usually required to conduct potentially uncertain, costly, or time-intensive studies in order to obtain FDA clearance. In contrast, competing manufacturers are often able to market similar devices within a year of De Novo clearance via the 510(k) process, which does not usually require premarket clinical evidence for clearance. To incentivize manufacturers to generate stronger evidence, the FDA could further collaborate with the Centers for Medicare and Medicaid Services (CMS) to align market clearance with payment policy. For example, the FDA and CMS could expand utilization of collaborative parallel premarket review programs to help expedite coverage for devices supported by compelling pivotal clinical data.^24,25^

The FDA and CMS could additionally leverage real-world data to evaluate these devices, which were largely cleared without comparative outcomes data. Key stakeholders in the medical device ecosystem – such as the FDA, payors, and manufacturers – are now developing the National Evaluation System for Health Technology (NEST) to enable timely real-world evidence generation to fill this gap.^26^ For example, NEST has funded a randomized clinical study of the effect of the Apple Watch electrocardiogram and irregular rhythm notification features (both cleared via the De Novo pathway in 2018) on patient-reported outcomes and clinical utilization (of note, this study is being conducted by SSD and JSR). More expeditious integration of Unique Device Identifiers into electronic data sources, particularly electronic health records and claims, would also facilitate real-world data based evaluations of De Novo devices.^17,27^ The European-based Institute for Quality and Efficiency in Healthcare and National Institute for Health and Care Excellence provide a model for the US to use real-world data to make recommendations that guide clinical practice. Data sharing and collaboration with such institutes could additionally strengthen patient protections against ineffective, unsafe, and low-value devices.

Our study has several limitations. First, we restricted analysis to therapeutic devices and our findings may therefore not be generalizable to all devices cleared via the De Novo pathway. Second, our analysis was cross-sectional. It is possible that additional postmarket studies will be required, more recalls will occur, and more 510(k) devices will be cleared for the devices in our sample. Third, our study is limited by the quality and availability of information within FDA review documents. Inaccuracies or omissions within these documents may have influenced our results.

In conclusion, the De Novo pathway has served as an increasingly important path to market for first-of-a-kind therapeutic devices. These devices rapidly serve as the basis for new models and competitor products. Although experience to date suggests that De Novo devices are largely safe, important questions about device effectiveness remain unanswered. Enhanced postmarket surveillance may enable the FDA to best utilize this pathway in the service of patients moving forward.

## Data Availability

All data used is available on public FDA databases, namely the De Novo database, the 510(k) clearance database, the 522 Post Market Surveillance database, and the Recalls of Medical Devices database

## Acknowledgements

VKR and JSR led study conception and design. JLJ contributed to study design and led drafting the manuscript, data acquisition, and analysis. The primary data sources were publicly available FDA documents. SSD, JSR, and VKR are subject matter experts in the regulation of medical devices. SSD and JSR have additionally received grant funding from the FDA and the Medical Devices Innovation Consortium, through the National Evaluation System for Health Technology, to pioneer methods of medical device postmarket surveillance and evaluation. JSR and VKR provided JLJ with supervision. All authors revised the manuscript for critically important content. VKR is the guarantor.

Mr. Johnston has received support from the FDA through the Yale-Mayo Clinic Center for Excellence in Regulatory Science and Innovation (CERSI) program. Dr. Dhruva currently receives research support through the National Institute of Health (K12HL138046) and the Greenwall Foundation. He also reports receiving travel support from the Food & Drug Administration and the National Evaluation System for health Technology Coordinating Center (NESTcc). In the past 36 months, Dr. Ross received research support through Yale University from Medtronic, Inc. and the Food and Drug Administration (FDA) to develop methods for postmarket surveillance of medical devices (U01FD004585), from the Centers of Medicare and Medicaid Services (CMS) to develop and maintain performance measures that are used for public reporting (HHSM-500-2013-13018I), and from the Blue Cross Blue Shield Association to better understand medical technology evaluation; Dr. Ross currently receives research support through Yale University from Johnson and Johnson to develop methods of clinical trial data sharing, from the Food and Drug Administration to establish Yale-Mayo Clinic Center for Excellence in Regulatory Science and Innovation (CERSI) program (U01FD005938), from the Medical Device Innovation Consortium as part of the National Evaluation System for Health Technology Coordinating Center (NESTcc), from the Agency for Healthcare Research and Quality (R01HS022882), from the National Heart, Lung and Blood Institute of the National Institutes of Health (NIH) (R01HS025164, R01HL144644), and from the Laura and John Arnold Foundation to establish the Good Pharma Scorecard at Bioethics International and to establish the Collaboration for Research Integrity and Transparency (CRIT) at Yale.

Dr. Rathi has no conflicts of interest or funding to disclose.

